# Development and cross-tissue validation of a methylation profile score for the cortisol response to stress

**DOI:** 10.64898/2026.02.17.26346504

**Authors:** David Balfour, Murthy Mittinty, Duc Phuc Nguyen, Sarah Cohen-Woods

## Abstract

Hypothalamic-pituitary-adrenal axis (HPA axis) dysregulation is a risk factor for poor mental and physical health. Animal studies indicate that DNA methylation may be one mechanism through which stress can influence the function of the HPA axis, however human studies have not identified consistent individual loci. Machine learning can be used to develop methylation profile scores (MPSs), but this method has not yet been applied to HPA axis function. Using a novel machine learning pipeline, we developed an MPS to predict the salivary cortisol response (AUCi) to the Trier Social Stress Test (TSST) from whole blood Illumina Infinium HumanMethylation 450K BeadChip data (*N* = 84, mean age = 34, 49% female). The MPS was associated with the cortisol response in an independent, cross-tissue cohort (*N* = 53, mean age = 20, 51% female), both before (β = 0.33, 95% CI [0.09, 0.54]) and after a social stressor (β = 0.3, 95% CI [0.09, 0.47]). Functional characterisation revealed several immune, stress, and disease-related pathways and genes, including tolerance induction to self antigen, chronic myeloid leukemia, *NR3C2*, and *PSMB4* (putatively causal in depression). We have developed and validated a novel epigenetic biomarker for stress reactivity, identifying a set of genomic loci where DNA methylation is associated with the cortisol response. Future research could investigate if HPA axis-related MPSs could be used alongside traditional risk factors to improve clinical risk assessment.

## Introduction

Stress can have a substantial impact on mental and physical health [1, 2, 3]. When exposed to a stressor, the hypothalamic-pituitary-adrenal axis (HPA axis) increases the level of cortisol, a hormone that facilitates widespread changes in immune function, metabolism, cognition, and brain function that prepare the individual to respond to a stressor. Prolonged exposure to an elevated level of cortisol due to chronic stress, or a physiological condition such as Cushing’s syndrome, can impact homeostasis across multiple bodily systems and predispose the individual to higher risk of poor mental and physical health outcomes, including depression, heart disease, and metabolic syndrome [4, 5, 6].

There are individual differences in the reactivity of the HPA axis, with some people showing a stronger or weaker cortisol response. These differences may have long-term effects on health, influencing vulnerability and resilience to stress [7]. The specific pathways that underpin differences in the function of the HPA axis are not well-understood. Twin studies on the cortisol response have reported heritability estimates of up to 45%, indicating a substantial genetic component [8]; at the same time, non-genetic factors appear to account for more than half of the observed variability. Studies in rodents suggest that differences in maternal care may lead to differences in DNA methylation at the gene *Nr3c1*, altering the hormonal response to stress [9, 10]. *Nr3c1* codes for the glucocorticoid receptor (GR), which facilitates the effect of cortisol, including negative feedback mechanisms that work to suppress the secretion of cortisol. DNA methylation is therefore a plausible mechanism through which the HPA axis could adapt to the environment, making it more – or less – reactive.

In humans, childhood trauma, and prenatal exposure to maternal stress, have been associated with HPA axis dysregulation and DNA methylation [11, 12, 13, 14]. The proposed relationship between DNA methylation and the function of the HPA axis is not as well-established. A recent systematic review and meta-analysis on DNA methylation and the cortisol response to an acute psychological stressor reported that findings in the literature are variable, although there is some tentative evidence for a relationship between *NR3C1* methylation and a stronger cortisol response in infants [15].

The statistical methods that have been used to date have not been sufficient to identify robust correlates of the cortisol response, with a majority of studies having been candidate gene studies or traditional methylome-wide association studies (MWAS) [15]. Candidate gene studies are known to produce unreliable results, so there is a broader move away from them toward more data-driven approaches such as MWAS [16, 17]. Unfortunately, MWAS require a high level of correction for multiple testing, which – with the small samples currently available – presents issues with statistical power. The largest MWAS for the cortisol response to date may not have had a power level above only 2.2% [15]. Another limitation of the existing research is that MWAS and traditional candidate gene studies usually investigate the relationship between individual genes or CpG sites, and the phenotype of interest, using separate linear models. However, as with other complex phenotypes, it is likely that the relationship between DNA methylation and the cortisol response depends on the joint effect of many different genomic loci, each of which has only a small effect in isolation.

Machine learning refers to the use of algorithms that can learn how to automate complex tasks that may be difficult to program manually [18], and has not yet been applied in this field. This method can provide new insight, as – unlike standard candidate gene and MWAS analyses – machine learning can simultaneously model the joint effect of multiple loci and uncover interactions and non-linear relationships in high-dimensional data. It can also detect subtle but genuine biological signals that would be missed after the correction for multiple testing in a standard MWAS. This can be done by focusing on predictive modeling instead of significance testing and using resampling (e.g., *k*-fold cross-validation), shrinkage, and validation using held-out test sets to mitigate overfitting. Machine learning methods have already been used to model the relationship between DNA methylation and a wide range of different phenotypes, including age, schizophrenia, and some proteins in blood plasma [19, 20, 21]. In each case, the resulting model is referred to as a methylation profile score (MPS) [22]. An MPS is a kind of epigenetic biomarker, providing a DNA methylation-based proxy or estimate of the phenotype of interest [22].

In this study, we aimed to generate a replicable MPS for the cortisol response to stress using a novel combination of machine learning techniques, including mutual information [23], lasso stability selection [24, 25], gradient boosting [26], and stacked ensembling [27]. These methods have been implemented separately in prior research, but they have not – to our knowledge – been brought together into a single, cohesive workflow. By developing the MPS, we aimed to identify and characterise genomic loci where DNA methylation may be jointly associated with the cortisol response, providing new insight into the relationship between DNA methylation and the HPA axis in humans.

## Methods

### Data collection

#### Training cohort

This study used publicly available data. The full training cohort (GSE77445) included 85 participants from the general population at the University Medical Center, Utrecht, The Netherlands [13]. The group Trier Social Stress Test (TSST-G), a group adaptation of the classic Trier Social Stress Test (TSST), was used to induce acute psychological stress. The TSST-G is a standard laboratory protocol involving a speech task and a mental arithmetic task in front of a panel of researchers and a video camera, conducted in groups of up to four [13, 28]. Saliva was sampled eight times over a 90-minute period, beginning approximately 10 minutes before the stressor. Area under the curve with respect to increase (AUCi) was used to represent the change in cortisol relative to the baseline value [29]. Whole blood was sampled before the test and DNA methylation was assayed using the Illumina Infinium HumanMethylation450K BeadChip.

#### Cross-tissue validation cohort

No datasets were available for same tissue validation. Validating across different tissue types presents a more stringent and challenging goal, so – as saliva DNA methylation data were publicly available – a cross-tissue validation approach was used. The full validation cohort included 55 undergraduate students recruited from the University of Texas at Austin [30]. An adaptation of the TSST, the Semi-Virtual Trier Social Stress Test (SV-TSST), was used to induce acute psychological stress. As in the TSST-G, participants were asked to complete a speech task and a mental arithmetic task in front of a panel, although it was conducted using video conferencing software (Zoom) and completed individually rather than in groups. Four saliva samples were obtained over an 80-minute period, beginning approximately 10 minutes before the stressor. As in the training cohort, AUCi was used to represent change in cortisol relative to the baseline value. In contrast to the training cohort, DNA methylation data was obtained both before and after the stressor. Also, DNA methylation was assayed in saliva, not blood, using the Illumina Infinium MethylationEPIC v1.0 BeadChip, a more recent version of the BeadChip used in the training cohort. DNA methylation data pre-processing steps are outlined in the supplementary methods. Informed consent and appropriate ethical approvals were obtained at the time of data collection, as detailed by the researchers who generated the data [13, 30].

### Model development

#### Overview

Several steps were used to develop an MPS for the cortisol response to stress (AUCi), as outlined in Fig. 1. Analyses were performed in R Version 4.5.1 [31].

**Fig. 1:**
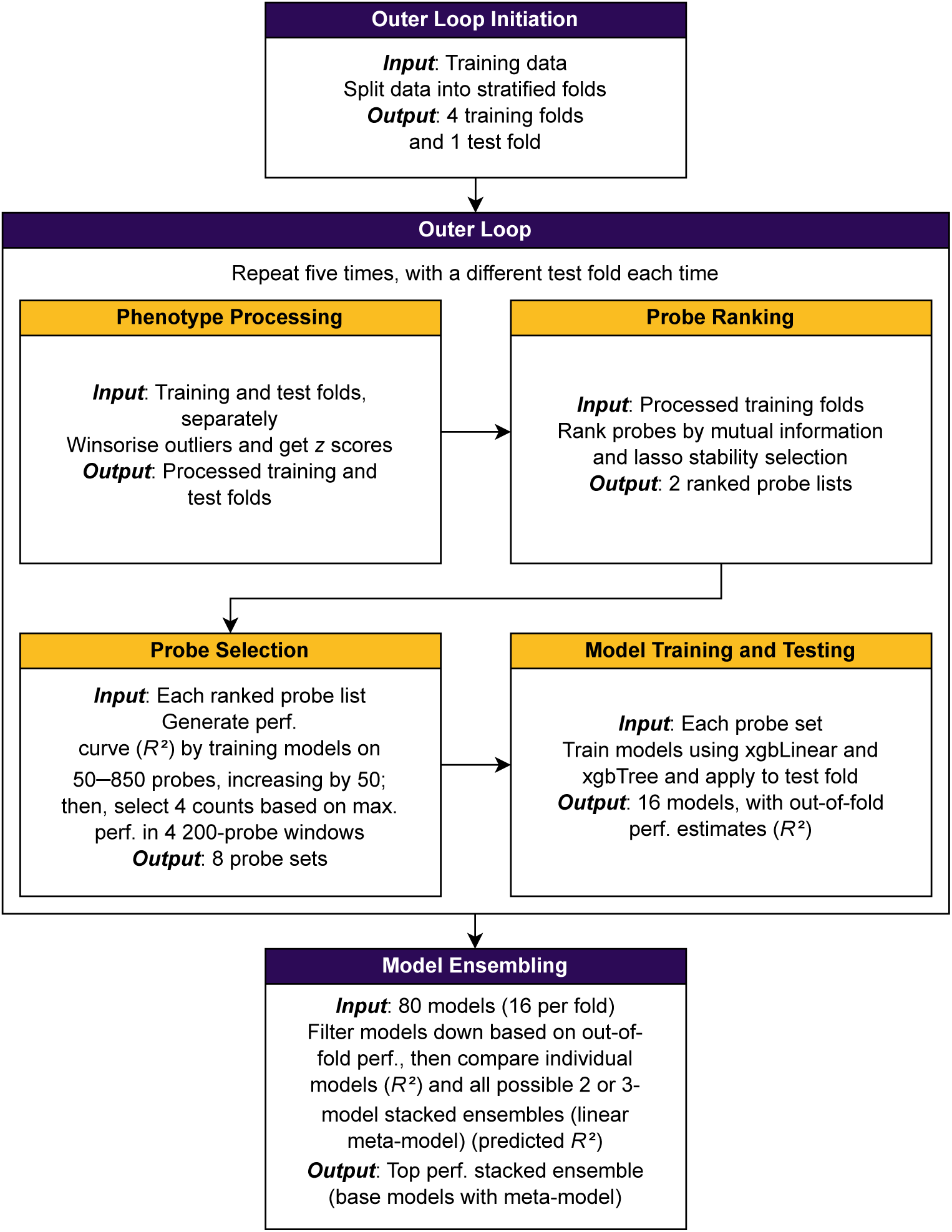
Outline of the process used to develop the MPS and obtain out-of-fold performance estimates in the training cohort.

In brief, we trained and tested a diverse selection of models using different machine learning techniques and sources of information and combined those that were the most effective in a held-out portion of the training cohort.

#### Outer loop

First, the training data were split into five stratified folds (groups), based on the cortisol response (AUCi). An outer loop was initiated, with five repeats using four folds for training and one for testing (i.e., an 80:20 split). Each repeat used a different fold for testing and included several steps, outlined below.

#### Phenotype processing

The cortisol response variable (AUCi) was processed separately in the training folds and the test fold to prevent data leakage. First, outliers were identified and winsorised using a 1.5 IQR threshold. The phenotype was then converted into *z* scores, standardized in order to account for the fact that measurements from different cortisol assays can differ substantially in absolute terms, which may limit their comparability [32, 33, 34].

#### Probe ranking

The probes were ranked using two different methods to identify the most potentially informative. The first method was mutual information, which indicates the dependency between two variables – in this case, the probe and the cortisol response [23, 35]. The second was lasso stability selection, a resampling technique for identifying the most important set of predictor variables [25]. First, the probes were filtered down to the top 850 based on their absolute correlation with the phenotype. Then half of the participants were randomly sampled 100 times. Each time, lasso – a penalised regression technique – was used to select a subset of probes to predict the cortisol response. The probes were then ranked by their probability of being selected across the iterations.

#### Probe selection

The next step, probe selection, involved selecting the number of probes to take forward for inclusion in the training data [35]. For each ranked probe list (mutual information or lasso stability selection), 17 probe sets were generated including the top 50 to 850 probes, increasing in units of 50. Correlation filtering was applied using the findCorrelation() function in the package *caret*, to reduce multicollinearity [36] (final probe counts in Supplementary Table 1). Performance curves were generated by training models to predict the phenotype with each filtered probe set. Following Doherty et al. [35], the models that were trained on the mutual information probe sets were generated with elastic net, a penalised regression technique. A tree-based algorithm, random forest, was used for the stability selection probe sets because elastic net would have been largely redundant, as it is lasso with an additional penalty. Four probe counts were selected based on maximum performance (*R*^2^) within four 200-probe windows. Windows were used instead of the overall maximum performance to encourage the selection and exploration of a wide range of probe counts.

#### Model training and testing

For each probe set, two models were trained, each using a different machine learning algorithm: xgbLinear or xgbTree. Both were implemented because it was not known at the outset which would be better in this context, and – also – because different algorithms among the base models can create diversity, leading to a more robust ensemble [37]. The xgbLinear model assumes the relationships of interest are linear, whereas xgbTree can model both linear and non-linear relationships, as well as interaction / moderation effects. Both algorithms use gradient boosting, a strategy where performance is gradually improved by training simple models (weak learners) one after the other, to correct the errors of a combination (ensemble) of the previous models [26]. Hyperparameters (settings that control the model’s behaviour during training) were tuned using a standard machine learning technique, repeated *k*-fold cross-validation, with three repeats and five folds. Out-of-fold *R*^2^ was obtained using the test fold (i.e., the fold that was left out of training for the current repeat of the outer loop).

#### Model ensembling

At the end of the outer loop, there were 80 different models, each representing a unique combination of fold, ranking method, probe count, and training method (5 folds x 2 ranking methods x 4 probe counts x 2 training methods). The models were filtered down to those with a positive out-of-fold *R*^2^, then a stacked ensemble [27] was created for each possible combination of two or three models. This involved using linear regression to fit a meta-model predicting the cortisol response from the predicted values of the selected models, referred to as base models. Predicted *R*^2^ was then computed for each ensemble, providing a less biased estimate of performance than *R*^2^ would when computed using the whole data set on which the model was fitted [38, 39]. The individual models were included in the comparison (using *R*^2^) to confirm that an ensemble would actually enhance performance. The ensemble with the strongest performance in the training cohort was taken forward as the MPS. A stacked ensemble (with models trained on different but overlapping subsets of the data) with gradient boosting was expected to optimise the bias-variance trade-off, with the former strategy working to reduce variance and the latter working to reduce bias.

#### Cross-tissue validation

The MPS was validated in an independent, cross-tissue cohort with publicly available data [30]. As in the training data, the cortisol response variable (AUCi) was winsorised using a threshold of 1.5 IQR, and converted into *z* scores. Linear regression models were fitted with the estimate from the MPS as the predictor and the processed cortisol response variable as the outcome. Initial analyses included no covariates to more closely match the conditions during training, but adjusted analyses were performed that included the following covariates: sex, age, self-reported ethnicity, and mCigarette (a recent smoking MPS [40]). Also, a fully-adjusted model was fitted that included all of the listed covariates plus immune cell proportion estimates (computed with the package *EpiDISH* [41], following Miller et al. [30]). Visual inspection of residual plots revealed potential deviations from normality, homoscedasticity, and linearity. To ensure valid inference despite potential assumption violations, 95% confidence intervals (CIs) were generated using non-parametric case bootstrapping with the adjusted bootstrap percentile method, as described by Fox and Weisberg [42, 43]. Statistical significance was determined based on whether the CIs included zero. Analyses used two-sided tests unless otherwise stated, and all reported regression coefficients are standardised.

#### SHAP analysis

We used SHAP analysis to help with the interpretation of the MPS. This involves generating SHAP values, which indicate – for each participant – how much each variable contributed to the predicted value [44]. As described in the supplementary methods, the SHAP values were used to obtain an importance score representing the percentage of the predictive signal contributed by each probe, averaged across the training and validation data. A subset of important probes (i.e., those with a non-zero mean importance score) were identified and taken forward for further analysis. For each data set, the potential_interactions() function in the package *shapviz* was used to obtain an estimate of the strength of the potential pairwise interaction or moderation effect for each probe with every other probe. These estimates were summed up to obtain a potential interaction / moderation score. The probes were ranked by their mean score across the data sets to identify those that may have a substantial effect in conjunction with other probes.

#### Functional characterisation

Several analyses were performed to characterise the probes that were identified as important based on their non-zero mean importance score across the data sets. The package *MethylToSNP* was used to screen the probes for a beta value distribution that may be indicative of possible bias due to an underlying SNP [45]. Empirical *p* values were generated using permutation tests to determine if the probes may be enriched or depleted for specific gene region feature or relation to (CpG) island annotations [46]. Gene set enrichment analysis was used on the set of genes that are annotated to the important probes. This was applied using the gometh() function in the package *missMethyl*, with the GO and KEGG databases and a false discovery rate (FDR) of .05. Also, the subset of important probes (i.e., those with a non-zero mean SHAP importance score) were examined for annotation to genes that were of particular interest due to prior research and known functional relevance: *CRH* [47], *CRHR1* [48], *FKBP5* [48], *HSD11B1* [49], *HSD11B2* [49], *LEP* [50] [51], *MC2R* [52], *NR3C1* [53], *NR3C2* [54], *OXTR* [55], *SKA2* [56], and *SLC6A4* [57].

Several analyses were performed to determine if the important probes may be sensitive to stress or glucocorticoids. Specifically, we investigated if any were previously identified as glucocorticoid-sensitive by Provençal [58]. Also, an empirical *p* value was generated using a permutation test to determine if the probes may be enriched or depleted for nominally stress-responsive probes [46], defined as those showing a nominally significant change from pre- to post-stress in the cross-tissue validation data using a paired *t*-test. A paired *t*-test was also used to investigate if the MPS changed from pre- to post-stress. Additional methodological details are available in the supplementary information.

#### Code availability

The code used in this study is available on request.

## Results

### Participant characteristics

Participant characteristics are reported in Table 1. The training cohort represented a wide age range from early adulthood to early old age and it was ethnically homogenous, with all participants being Caucasian. Participants in the validation cohort were all in early adulthood and they were more ethnically diverse, with Asian / Pacific Islander being the most common demographic. Cortisol AUCi was higher in the training group, possibly due to different cortisol assays and measurement procedures [33, 34].

### Model performance in the training cohort

The top-performing model in the training cohort was an ensemble that included three base models: one trained using xgbLinear (mutual information, 650 probes) and two trained using xgbTree (lasso stability selection, 349 probes; lasso stability selection, 200 probes). These models achieved out-of-fold *R*^2^ values of .27, .09, and .08 (Fig. S1) (base model hyperparameters and tuning plots: Supplementary Table 2 and Fig. S2 to S4; ensemble weights: Supplementary Table 3).

### Independent cross-tissue validation

#### Unadjusted analyses

The final MPS (i.e., the ensemble) was associated with the cortisol response in the cross-tissue validation cohort, both before and after the stressor (before: *R*^2^ = .11, β = 0.33, 95% CI [0.09, 0.54], Fig. 2; after: *R*^2^ = .09, β = 0.3, 95% CI [0.09, 0.47], Fig. 3).

**Fig. 2:**
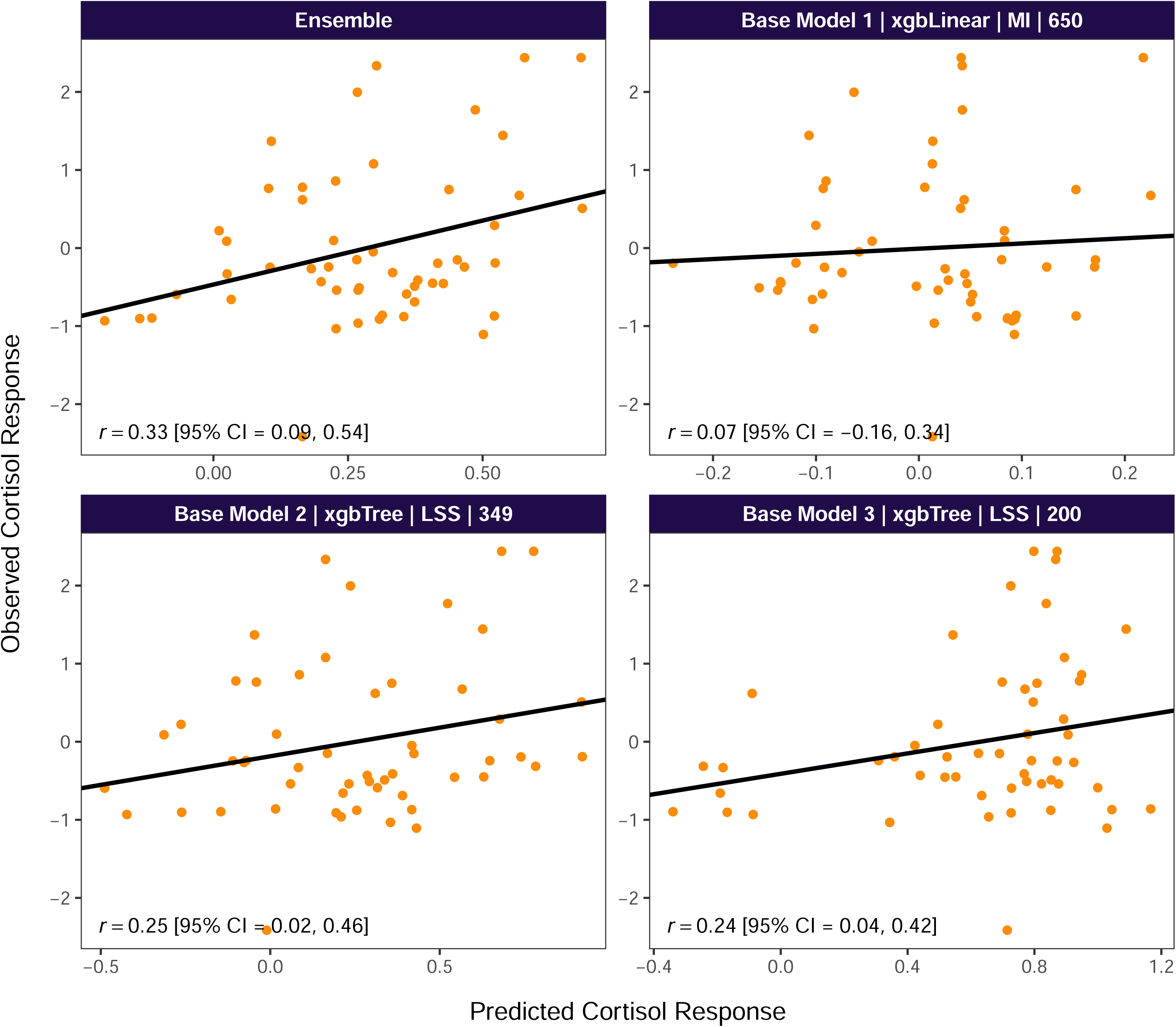
Pre-stress performance of the MPS in the cross-tissue validation cohort. Unadjusted correlation between observed winsorised AUCi z-scores and the MPS scores (which were trained on winsorised AUCi z-scores). MI = mutual information probe ranking, LSS = lasso stability selection probe ranking. Number in facet title indicates the number of probes in the model.

**Fig. 3:**
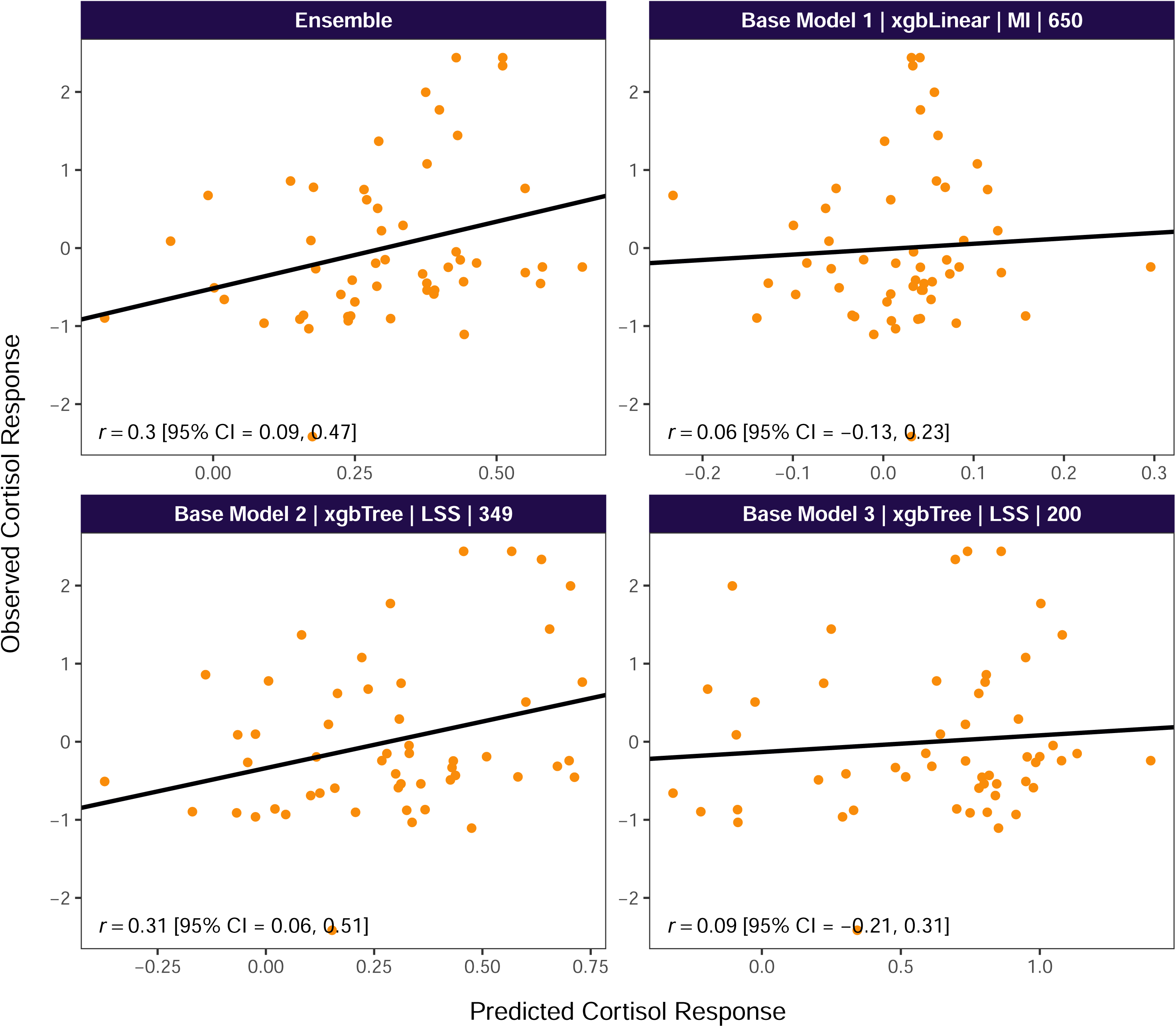
Post-stress performance of the MPS in the cross-tissue validation cohort. Unadjusted correlation between observed winsorised AUCi z-scores and the MPS scores (which were trained on winsorised AUCi z-scores). MI = mutual information probe ranking, LSS = lasso stability selection probe ranking. Number in facet title indicates the number of probes in the model.

#### Adjusted analyses

The effect of the MPS remained statistically significant after adjusting for age, sex, ethnicity, and smoking (pre-stress: β = 0.27, 95% CI [0.002, 0.50]; post-stress: β = 0.32, 95% CI [0.08, 0.53]). When cell proportion estimates were added to the model, effects became non-significant in the pre-stress data, but remained significant in the post-stress data (Table 2).

In the fully-adjusted model, Non-Hispanic White ethnicity and cell proportion estimates were significant independent predictors of the cortisol response (Table 2). In models that only adjusted for ethnicity or the cell proportion estimates without the other covariates, the effect of the MPS remained significant (ethnicity, pre-stress: 0.32, 95% CI [0.08, 0.53], ethnicity, post-stress: 0.35, 95% CI [0.11, 0.53]; cell proportion estimates, pre-stress: 0.29, 95% CI [0.05, 0.50], cell proportion estimates, post-stress: 0.28, 95% CI [0.1, 0.47]). The MPS was not significantly associated with ethnicity, total leukocyte proportion, the neutrophil-to-lymphocyte ratio [59], or any of the individual cell proportion estimates, in the training data or in the validation data, pre- or post-stress (*p* > .05).

#### SHAP analysis

There were 1 171 unique probes in the ensemble (Supplementary Table 4). Of those, 425 had a non-zero mean SHAP importance score across the data sets (Supplementary Table 5). The most important probes were cg20174000 (*ABL1*), cg03341991 (*N4BP2L2*), and cg27224971 (not annotated to a protein-coding gene) (Fig. 4).

**Fig. 4:**
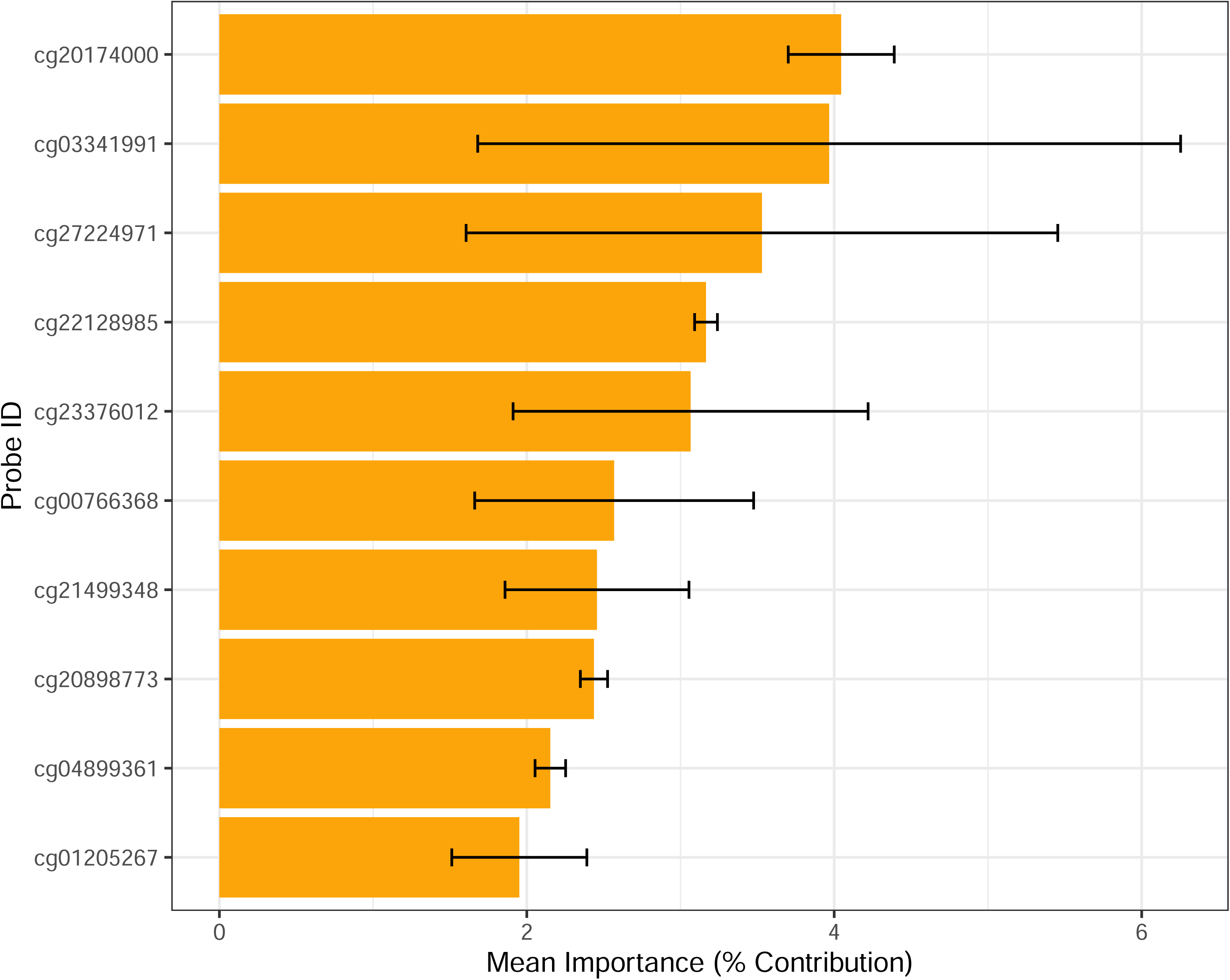
Top MPS probes by mean SHAP importance across the data sets. Bars show mean SHAP importance score across the data sets, representing the percentage of the predictive signal contributed by the probe. Errors bars show the standard deviation.

The probes with the highest mean interaction / moderation score across the data sets were cg22736107 (*PSMB4*), cg05541125 (*ATP5O*), and cg14681854 (*EVI5*) (Supplementary Table 6).

#### Functional characterisation

There was no evidence to suggest the important probes were biased by underlying genetic variance, using *MethylToSNP*. Two-sided permutation tests revealed some evidence to suggest the important probes may be overrepresented in the north shores of CpG islands (observed: 16.47%, null: 12.98%, *p* = .03) and underrepresented within 200 bases of the transcription start site (observed: 10.12%, null: 13.37%, *p* = .05) (Supplementary Table 7). None of the important probes were previously identified by Provençal [58] as glucocorticoid-sensitive. Also, there was no evidence for enrichment or depletion for nominally stress-responsive probes (i.e., probes that changed significantly from pre- to post-stress) (observed: 26.35%, null: 26.98%, *p* = .79). The MPS did not change significantly from pre- to post-stress, paired *t*(52) = -0.57, *p* = .57. One of the important probes (cg05075176) was annotated to a gene of prior interest, *NR3C2*, in the north shore of a CpG island. Gene set enrichment analysis revealed no significant GO terms or KEGG pathways at a FDR < .05 (Supplementary Table 8 and Supplementary Table 9). The GO terms with the lowest uncorrected *p* values were tolerance induction to self antigen (*p* = .000005), leukocyte homeostasis (*p* = .0002), and dendritic spine organization (*p* = .0002). The KEGG pathways with the lowest uncorrected *p* values were primary immunodeficiency (*p* = .003), chronic myeloid leukemia (*p* = .01), and endocrine resistance (*p* = .01).

## Discussion

We developed a blood-based MPS for the cortisol response to stress that replicated in an independent cohort in saliva-derived DNA. The MPS was significantly associated with the cortisol response after adjustment for smoking and standard demographic variables, suggesting it is not likely to be confounded by them. Additional adjustment for estimated immune cell proportions attenuated the pre-stress relationship, suggesting the MPS may in part represent differences in cell composition associated with the cortisol response. That would help to explain the cross-tissue replication, as immune cells have distinct epigenetic signatures that are present in both blood and saliva. At the same time, several findings are consistent with a predictive signal beyond cell composition. In particular, the MPS remained a significant predictor of the cortisol response in a fully-adjusted model at the post-stress time point. Also, it was significantly associated with the cortisol response in a model that adjusted for the cell proportion estimates without the other covariates, both before and after the stressor. Moreover, it was not associated with the total proportion of leukocytes or the individual cell proportion estimates. Together, these findings indicate that although the MPS may capture some variance associated with immune cell proportions, it does not appear to be a simple representation of them. Like other MPS that have been developed using peripheral tissue, it may represent a combination of between- and within-cell variance in DNA methylation associated with the phenotype [22].

The results of the functional characterisation are consistent with the possibility that the MPS may capture immune-related variance. Although there were no significant results for gene set enrichment analysis after correcting for multiple testing, the top nominally significant GO term was tolerance induction to self antigen. This refers to the process through which the immune system differentiates between self and non-self antigens, preventing autoimmunity [60]. The second GO term was leukocyte homeostasis, which refers to the process through which the body maintains a stable balance of immune cells. The top KEGG pathways were primary immunodeficiency (genetically determined immune disorders) and chronic myeloid leukemia, a cancer involving the malignant expansion of haematopoetic stem cells [61]. The probe with the strongest contribution to the predicted value (i.e., the highest SHAP importance) was annotated to *ABL1*, an oncogene involved in the development of chronic myeloid leukemia [62]. The probe with the second highest contribution was annotated to *N4BP2L2*, a gene that may play a role in neutropenia (i.e., a deficiency of the most abundant type of white blood cell) [63]. When the probes were ranked based on the sum of their potential interaction or moderation effects, the top probe was annotated to *PSMB4*. This gene codes for a subunit of the proteasome, a protein complex involved in the maintenance of protein homeostasis [64]. A specialised form of the proteasome, the immunoproteasome, is involved in the processing of class I major histocompatibility complex peptides, which are presented on the surface of the cell for recognition by T cells [64, 65]. Recently, a Mendelian randomisation study reported evidence to suggest that *PSMB4* may play a causal role in depression [66]. Overall, the MPS appears to be associated with immune and disease-related pathways and genes.

It is possible that the MPS, while designed to represent stress reactivity, is simultaneously capturing, to some extent, the effect of chronic stress on DNA methylation, the immune system, and the function of the HPA axis [67, 68]. The MPS does not appear to be sensitive to acute stress, as it did not change significantly from pre- to post-stress, and there was no evidence of enrichment for stress-responsive probes or probes that were previously identified as glucocorticoid-sensitive (i.e., those that may be sensitive to stress through the effect of cortisol). At the same time, as noted above, it appears to contain a predictive signal beyond cell composition. Longitudinal studies that investigate if the MPS may change in response to chronic or acute but extreme stress (e.g., combat, poverty, the start of a school term) would be informative. The MPS could also be investigated in people with leukemia, potentially providing new insight into the molecular pathology underpinning cancer-related HPA axis dysregulation [67, 69].

While speculative, it is possible that the MPS may capture, in part, the regulatory effect of DNA methylation on the cortisol response. Consistent with a regulatory role for some of the important probes, there was some evidence to suggest they may be overrepresented in the north shores of CpG islands, CpG-rich sequences of DNA that can function as genomic platforms for regulating transcription at associated promoters [70]. Also, one of the important probes is located in the north shore of a CpG island associated with the gene *NR3C2*, which codes for the mineralocorticoid receptor (MR). Like the GR, the MR facilitates the effect of cortisol, including negative feedback [71]. Contrary to research in rodents and popular theoretical models, none of the probes in the MPS are annotated to *NR3C1*. This finding is consistent with prior research, including a recent meta-analysis that revealed no evidence for an association between *NR3C1* DNA methylation and the cortisol response to an acute psychological stressor in adults [15]. Also, a candidate gene study reported evidence for a relationship between *NR3C2* methylation but not *NR3C1* methylation and the cortisol response to stress [54]. Consistent with a regulatory role for *NR3C2* but not *NR3C1* in adults, a recent experimental study reported that pharmacological blockade of the MR but not the GR resulted in an altered cortisol response to the TSST [71]. At the same time, there has been some mixed evidence for a genetic effect of *NR3C1* on the cortisol response [72, 73]. It may be that *NR3C2* dynamically regulates the cortisol response, potentially through epigenetic mechanisms, while *NR3C1* has a more static and largely genetically determined role. Also, time and the severity of the stressor may be relevant.

Although it is well-established that the GR mediates negative feedback within the HPA axis, it may be that its effect unfolds over a longer period of time or in response to a more severe stressor [71]. The MR, by contrast, may regulate the shorter-term response to a relatively mild stressor such as the TSST [71]. Overall, our findings fit into an emerging picture of *NR3C2*, but potentially not *NR3C1*, as a dynamic regulatory mechanism underpinning the cortisol response to an acute psychological stressor in human adults. Although our findings only shed light on the role of these mechanisms in peripheral tissue, they are consistent with the pharmacological blockade study cited above, which used antagonists that cross the blood-brain barrier [71].

Methodologically, this study shows promise for a novel combination of machine learning techniques for the development and interpretation of MPS, including mutual information [23], lasso stability selection [25], gradient boosting [26], stacked ensembling [27], and SHAP analysis [44]. The benefit of the stacked ensemble can be seen in the fact that it outperformed the base models both in the training data and in the pre- and post-stress validation data (Fig. 2 and Fig. 3). Moreover, this study provides a clear demonstration of how SHAP analysis can be used to identify the most important probes in a complex MPS, providing a focus for subsequent functional characterisation. Similar methods could be applied to other psychiatric and stress-related phenotypes in future. Research could also investigate if HPA axis-related MPSs could be used alongside traditional risk factors to improve clinical risk assessment.

A limitation of this study, shared by the majority of research to date on the cortisol response in humans, is the use of peripheral tissue. While peripheral tissue is relevant to the cortisol response, as this phenotype is measured in blood or saliva, it is not as theoretically relevant as other tissue types that play a more direct role in the function of the HPA axis (e.g., the hippocampus, the hypothalamus, the pituitary gland, and the adrenal glands). Unfortunately, it may not be feasible to obtain contemporaneous data on the cortisol response and DNA methylation in these tissues. Although we observed no evidence for an effect of *NR3C1* methylation in saliva and blood, it is possible that – as observed in rodents – hippocampal DNA methylation at this gene does regulate the hormonal stress response. Although data on stress reactivity are not, to our knowledge, available in conjunction with data on DNA methylation from human brain tissue, existing hippocampal samples (e.g., from the NIMH Human Brain Collection Core, [74]) could be leveraged to investigate if the MPS is associated with psychiatric or stress-related conditions. Finally, the sample size in this study is small considering the high-dimensional data, and – although the use of cross-validation and other techniques appear to have mitigated the problem of high-dimensionality, enabling successful replication – this study should be considered a proof of principle and a demonstration of a methodology that may be fruitfully applied to larger cohorts in future, pending the availability of appropriate data.

We developed and validated a novel epigenetic biomarker for stress reactivity. In doing so, we identified a set of genomic loci that may be jointly associated with the cortisol response to stress, providing new insight into the relationship between DNA methylation and the HPA axis in humans. The MPS may capture some variance associated with immune cell proportions, although it does not appear to be a simple representation of them. Functional characterisation revealed several immune, stress, and disease-related pathways and genes, including a putatively causal locus in depression (*PSMB4*). The genes that are annotated to the probes in the model also include *NR3C2* but not *NR3C1*, consistent with emerging evidence for the former, but potentially not the latter, as a dynamic regulatory mechanism underpinning the cortisol response to a relatively mild acute psychological stressor in human adults. Methodologically, this study provides a clear proof of principle for a novel combination of machine learning techniques for the development and interpretation of MPS. Future research may provide additional insight by investigating if the developed MPS is associated with longitudinal stress exposures and related psychiatric disorders in brain tissue. Studies could also investigate if HPA axis-related MPSs could be used alongside traditional risk factors to improve clinical risk assessment.

## Data Availability

Study data are available at the following links: training cohort (https://www.ncbi.nlm.nih.gov/geo/query/acc.cgi?acc=GSE77445), cross-tissue validation cohort (https://github.com/fchampagneUT/AcuteStressMethylation).

https://github.com/fchampagneUT/AcuteStressMethylation

## Acknowledgements

Thank you to the researchers who generously uploaded their data to public repositories, enabling the present study. DB’s work on this study was supported by an Australian Government Research Training Program Scholarship.

## Conflict of interest

The authors have nothing to disclose.

### Supplementary information

Supplementary information is available at MP’s website. supplementary_tables.xlsx.

Supplementary Tables 1 to 9. supplementary_information.pdf. Supplementary methods and Fig. S1 to S4.

Supplementary Table 1. Probe counts before and after correlation filtering.

Supplementary Table 2. Selected base model hyperparameters.

Supplementary Table 3. Meta-model weights.

Supplementary Table 4. Annotation for the unique probes in the ensemble model.

Supplementary Table 5. Mean SHAP importance score across the data sets.

Supplementary Table 6. Mean interaction / moderation score across the data sets.

Supplementary Table 7. Results of permutation tests for enrichment or depletion for relation to island and gene region feature annotations.

Supplementary Table 8. Results of gene set enrichment analysis using GO.

Supplementary Table 9. Results of gene set enrichment analysis using KEGG.

