## Supplementary Information for "Development and cross-tissue validation of a methylation profile score for the cortisol response to stress"

David Balfour^a^, Murthy Mittinty^b^, Duc Phuc Nguyen^c^, Sarah Cohen-Woods^a, d, e^

^a^College of Education, Psychology and Social Work, Flinders University, Bedford Park, South Australia, Australia

^b^College of Medicine and Public Health, Flinders University, Bedford Park, South Australia, Australia

^c^Flinders Health and Medical Research Institute – Sleep Health (Adelaide Institute for Sleep Health), College of Medicine and Public Health, Flinders University, Bedford Park, South Australia, Australia

^d^Flinders Centre for Innovation in Cancer, Bedford Park, South Australia, Australia

^e^Flinders University Institute for Mental Health and Wellbeing, Flinders University, Bedford Park, South Australia, Australia

Correspondence concerning this article should be addressed to:

David Balfour, PO Box 307, Crafers, South Australia, 5152, Australia. Phone: +61 413 157 032

Professor Sarah Cohen-Woods, Flinders University Institute for Mental Health and Wellbeing, Flinders University, Sturt Road, Bedford Park, South Australia, 5042, Australia. Phone: +61 468 437 245

### Supplementary methods

#### Data collection

##### Training cohort

Exclusion criteria included prescription medication, previous participation in stress-related research, and the presence of a mental or physical disorder based on the Mini-International Neuropsychiatric Interview (MINI) [1]. Saliva samples were obtained with Salivettes. Cortisol was measured using an in-house radioimmunoassay. DNA was extracted from whole blood using the Gentra Puregene kit (Qiagen, Valencia, CA, USA) and bisulphite conversion was performed using Zymo kits (Zymo Research, Orange, CA, USA).

##### Cross-tissue validation cohort

Exclusion criteria included substance abuse, nicotine use, an irregular menstrual cycle, pregnancy, breastfeeding, and the use of endocrine-related medications (excepting hormonal contraceptives) [2]. Saliva was obtained for cortisol analysis by asking participants to drool through a straw into a tube. Cortisol concentrations were assayed using a high throughput liquid chromatography-tandem mass spectrometry assay (Dresden LabService GmbH). Saliva samples were obtained for DNA methylation analysis using the Oragene-500 collection kit (DNA Genotek Inc.), which involves drooling into a tube. DNA was extracted using the MagMAX™ DNA Multi-Sample Ultra 2.0 Kit (Thermo Fisher Scientific) and the KingFisher™ Flex System. Bisulphite conversion was performed using the EZ-96 DNA Methylation™ Kit (Zymo Research).

#### DNA methylation data pre-processing

Prior to this study, several DNA methylation pre-processing steps were implemented by the researchers who published the data [1, 2]. In the training cohort, probe design biases were removed using Beta MIxture Quantile dilation (BMIQ) normalisation, and batch effects (Sentrix array and position) were removed using ComBat. Probes were removed if they had a detection *p* value > .001 and a bead count < 5 or if they were within 10 base pairs of the primer and at a SNP with a minor allele frequency > 5%. In the cross-tissue validation cohort, background correction was performed using the noob method. Probes were removed if ≥ 5% of samples had a detection p value > . 001 or a bead number < 4. Cross-reactive and gap probes were also removed and ComBat was used to remove batch effects. For the present study, outlier samples were screened for using the first two DNA methylation principal components [3] and a *z* score threshold of three. One outlier was removed from the training cohort and two were removed from the validation cohort, resulting sample sizes of 84 and 53, respectively. Beta values were converted to M values, which more closely approximate a normal distribution [4].

#### Model development

##### Outer loop

The repeats in the outer loop mitigated the risk of a single train/test split that would – by chance – be unsuitable for predicting the phenotype. The repeats also created diversity among the models, as the models were generated with five partially distinct data sets.

##### Phenotype processing

As noted in the main text, the phenotype was standardised because cortisol assays can differ subtantially in terms of the absolute values they produce. In some cases, this can yield values that differ by a factor of three or more [5, 6]. It is therefore not advisable to use cortisol values from different assays or laboratories in the same analyses without standardisation [5, 6, 7].

##### Probe ranking

Mutual information was computed for each probe in relation to the cortisol response using the mutinformation() function in the package *infotheo*. Lasso [8] was applied using the glmnet method in the package *caret*, with the alpha parameter set to 1. Lasso can be particularly useful when working with high-dimensional data because it reduces the coefficients of some of the predictors to 0, effectively selecting those that were assigned a non-zero coefficient.

##### Probe selection

In each probe set, to reduce multicollinearity, pairs of probes with a correlation (*r*) between them greater than .9 were identified [9]; then, the probe with the lowest correlation with the cortisol response (AUCi) was removed, as it was less likely to be informative regarding the outcome [9] (probe counts before and after filtering in Supplementary Table 1). This procedure was implemented using the findCorrelation() function in the package *caret*. Elastic net was implemented using the glmnet method in *caret*. Random forest was implemented in the same package, using the ranger method. As noted in the main text, random forest was used for the stability selection probes because elastic net would have been largely redundant, as it is lasso with an additional penalty. Also, the use of random forest at this point in the pipeline has the benefit of increasing diversity, drawing on multiple techniques sequentially to help identify the most relevant probes (in this case, a tree-based model and a model based on linear regression). As noted in the main text, probe counts were selected based on performance within windows rather than across the entire curve. This encouraged the selection and exploration of a wider range of probe counts. It also reduced potential redundancy, as the best-performing counts were often close to one another (e.g., 100 and 150).

##### Model training and testing

The xgbLinear and xgbTree models were trained with the package *caret*. The specific combination of these two models was expected to optimise the bias-variance trade-off. xgbLinear is expected to have lower variance (potentially at the cost of higher bias) because it assumes the relationships of interest are linear, whereas xgbTree is expected to have lower bias (potentially at the cost of higher variance) because it makes no such assumption – as it is a tree-based model, it can model both linear and non-linear relationships, as well as interaction / moderation effects.

Machine learning algorithms have hyperparameters, which are parameters outside the model itself that influence the behaviour of the algorithm during training. It is not usually possible to know at the outset which settings are ideal, so we used a standard technique, repeated *k*-fold cross-validation, to select them for each model. This involved splitting the training data into five folds (groups), then training models with different hyperparameter settings on four of the folds and testing them on the fifth fold. This was repeated five times, using a different fold for testing each time. The entire process was repeated three times, using a different random fold split each time. Model performance (*R*^2^) was averaged across the folds and repeats to identify the most effective hyperparameter settings. The final model was trained on the full training set with those settings. The tuneLength argument in the train() function in *caret* was used to automatically generate a suitable grid of hyperparameter values to explore.

##### Model ensembling

As explained in the main text, the models were filtered down to those with a positive out-of-fold *R*^2^. *R*^2^ was calculated as 1 - residual sum of squares / total sum of squares, in contrast with the default metric referred to as *R*^2^ in the *caret* package (Pearson’s *r*^2^), which is always positive and can mask poorly-performing models. The subsequent ensemble step involved generating new predictions for the base models on the whole training data set, to make the best use of the limited available data, then regressing the cortisol response variable on the predicted values. Predicted *R*^2^ was used to compare all possible two or three-model ensembles. Predicted *R*^2^ is computed using the sum of squares of residuals obtained from fitting the model *n* times, on *n* - 1 participants (also known as the predicted residual error sum of squares, or PRESS), providing a less biased estimate of model performance than *R*^2^ [10, 11]. The results of the procedure are mathematically equivalent to leave-one-out cross-validation (LOOCV) [12]. As noted in the main text, the individual models were included in the ensemble comparison to ensure an ensemble would actually provide a performance improvement. *R*^2^ rather than predicted *R*^2^ was used for the individual models, so as to not disadvantage them by fitting a redundant single-predictor meta-model. We acknowledge that comparisons using *R*^2^ and predicted *R*^2^ may have favoured xgbTree due to its lower bias compared to xgbLinear. However, *R*^2^ and predicted *R*^2^ were nevertheless used as standard metrics for performance on a regression task. The intercept and the slopes from the strongest ensemble (highest predicted *R*^2^) were saved, being the weights in a meta-model that could subsequently be used to generate predictions from the predictions of the base models.

#### Cross-tissue validation

The predicted values were generated from the three base models and combined into a final estimate using the meta-model. Some of the model probes were not available in the validation cohort due to differences in array design between the 450K and EPIC platforms (7.26%) or the quality control pipeline (1.71%). To bridge the gap between platforms without introducing data leakage, missing values were imputed using probe-wise medians from an independent, external saliva cohort (GSE119078, *n* = 28 healthy controls) [13]. This data set was selected because it used the required platform (i.e., the Illumina Infinium HumanMethylation 450K BeadChip, which has the majority of the missing probes) and the same tissue type as the validation cohort (saliva). Because the probes were completely missing (largely due to array design), more complex techniques that leverage available observations in the target data, such as *k*-nearest neighbours (*k*-NN), were not applicable. By substituting representative median values from a healthy saliva cohort, the chosen strategy neutralised the variance of the missing loci and provided a biologically plausible, tissue-specific baseline.

Immune cell proportion estimates were included in the fully-adjusted regression models. These were obtained using the hierarchical function hepidish() in the package *EpiDISH* with both an epithelial reference (centEpiFibIC.m) and the recommended immune cell reference in the package documentation (centBloodSub.m). The following cell types were included in the fully-adjusted models: B cells, NK cells, CD4T^+^ cells, monocytes, and granulocytes. Estimates for CD8T^+^ cells were omitted, as they were uniformly zero. As there were only three participants in the ‘Other’ ethnicity for the validation data, to ensure model stability, the ‘Other’ category was merged with the next-smallest category, ‘Hispanic White’.

As noted in the main text, visual inspection of residual plots revealed potential deviations from normality, homoscedasticity, and linearity. A quadratic polynomial term was added to accommodate the potential non-linearity, but this was abandoned as it failed to resolve the patterns observed in the residuals and introduced multiple influential points with Cook’s distance > 0.5. Potential interactions were also systematically investigated. These were not included in the final model, as none were significant at *p* < .05. Non-parametric case bootstrapping with the adjusted bootstrap percentile method [14, 15] was used to ensure valid inference despite potential violations of assumptions, as described in the main text. Variance inflation factors were all below five except for the cell proportion estimates, which were highly collinear, although this was not regarded as a problem as it could have no effect on the inferences drawn regarding the predictor of interest (i.e., the MPS), which was not collinear with the other variables.

#### SHAP analysis

As explained in the main text, SHAP analysis was used to help with the interpretation of the MPS. SHAP analysis involves generating SHAP values, which indicate – for each participant – how much each variable contributed to their predicted value [16]. Several steps were taken to rank the probes in the MPS by their SHAP values across the training cohort and the pre-stress and post-stress data in the cross-tissue validation cohort. First, to ensure comparability between the data sets, the probes were filtered down to those that were available in all three. Then, within each data set, raw SHAP values were generated for each base model using the package *fastshap* with the adjust parameter set to TRUE so the values for each participant would sum up to the difference between the participant’s predicted value and a given baseline value. Following the *fastshap* documentation, the baseline value was set to the mean predicted value in the training data. This was done by using the training data for the relevant base model as the background data, meaning the raw SHAP values represent the extent to which the probes pushed the predicted value above or below the average predicted value in the data the model were trained on.

To obtain ensemble SHAP values, the raw SHAP values for the base models were multiplied by the relevant coefficients in the meta-model and then added up. To obtain a standardised, data set-level representation of probe importance, the mean absolute ensemble SHAP value for each probe was divided by the sum of all mean absolute ensemble SHAP values in that data set and multiplied by 100. This resulted in a score representing the percentage of the predictive signal that was contributed by each probe. The mean and standard deviation were obtained for the percentage scores across the data sets to obtain an overall idea of importance. A subset of important model probes were then identified, defined as those with a non-zero mean importance score. Subsequent functional characterisation focused on this probe set, as there was no evidence that probes with a score of zero were used by the model, although they were included in the training data.

#### Functional characterisation

As described in the main text, permutation tests were used to determine if the probes in the MPS may be enriched or depleted for particular gene region feature and relation to island annotations. The following gene region feature annotations were tested: 3' UTR, 5' UTR, 1^st^ exon, body, TSS 200, TSS 1500. The following relation to island annotations were tested: island, north shore, south shore, open sea. For each test, an empirical null distribution was generated using 1 000 permutations of random sampling from the probes in the training data. Empirical *p* values were calculated using the formula (*b* + 1) / (*m* + 1) [17], where *m* was the total number of permutations (1 000). For one-sided *p* values, *b* was the number of permutations where the proportion of probes with the annotation was equal to or more extreme than the observed proportion. For two-sided *p* values, *b* was the number of permutations where the absolute deviation of the permuted proportion from the mean of the null distribution was greater than or equal to the absolute deviation of the observed proportion from the same mean. This approach was also used to test for enrichment or depletion for nominally stress-responsive probes.

### Supplementary figures

Fig. S1: Top models in the training cohort by out-of-fold performance

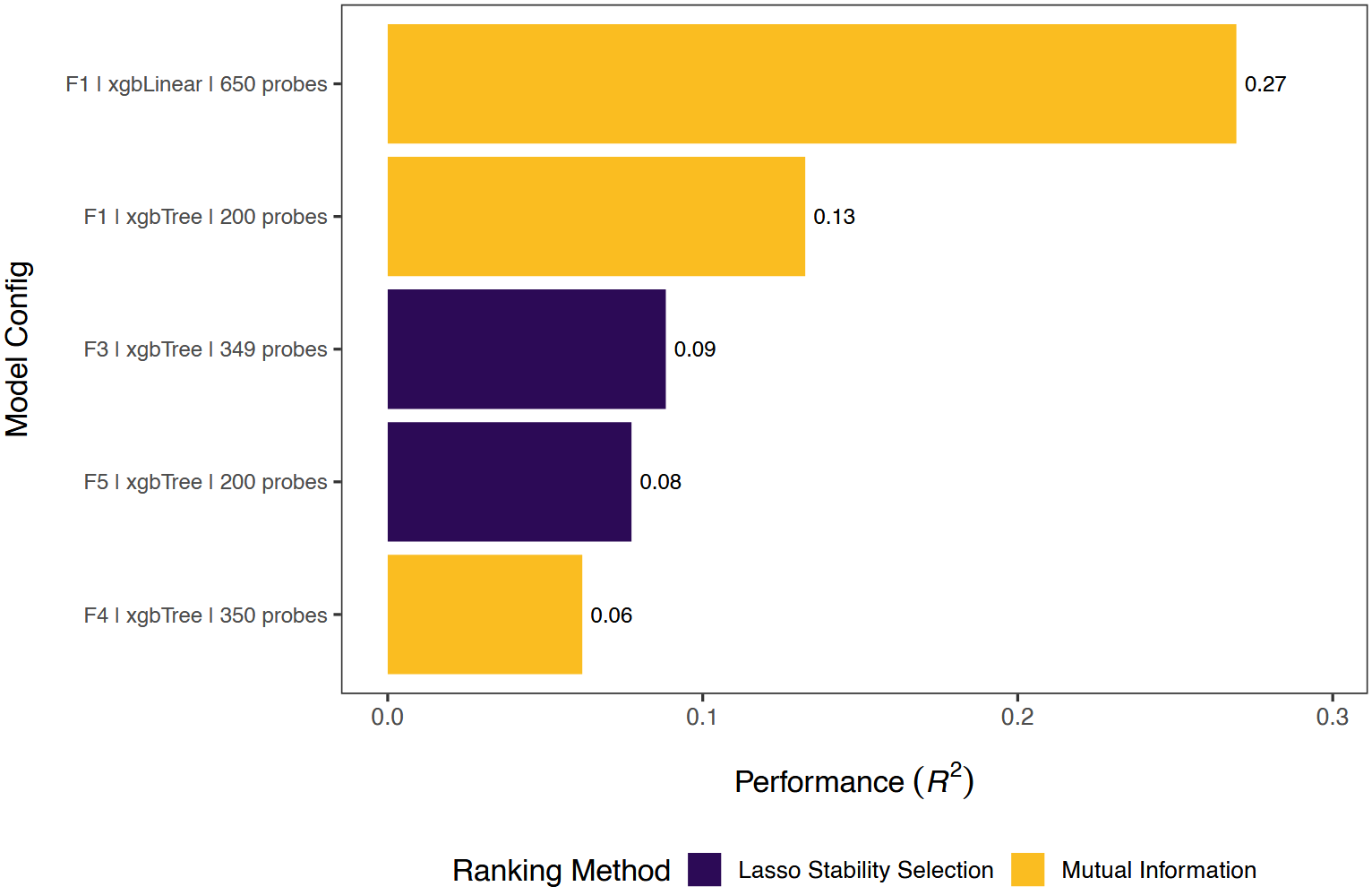

In the labels on the x axis, ‘F’ refers to the fold of the outer loop that was used for model testing (with the other folds being used for training).

Fig. S2: Tuning plot for base model 1

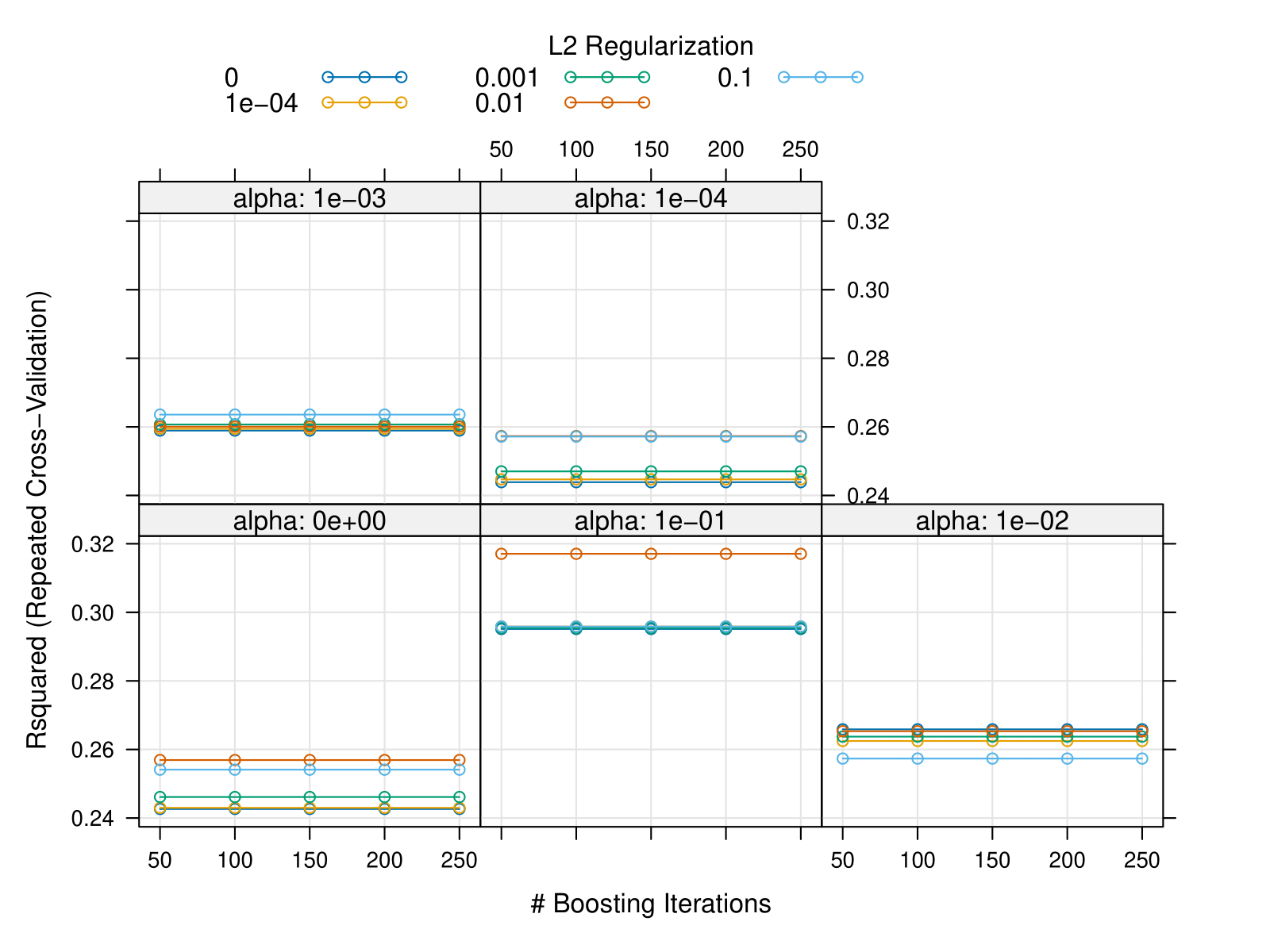

Fig. S3: Tuning plot for base model 2

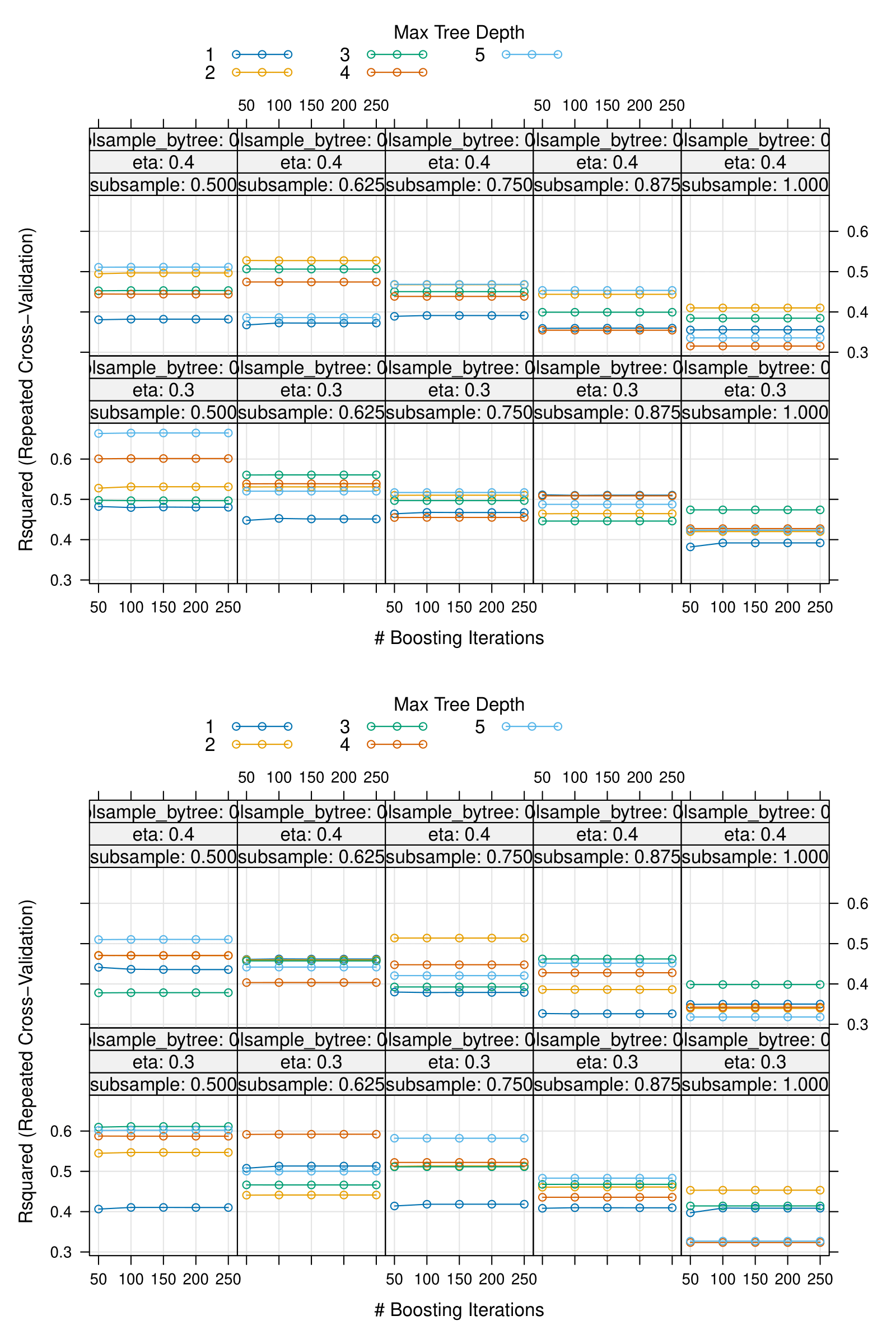

Fig. S4: Tuning plot for base model 3

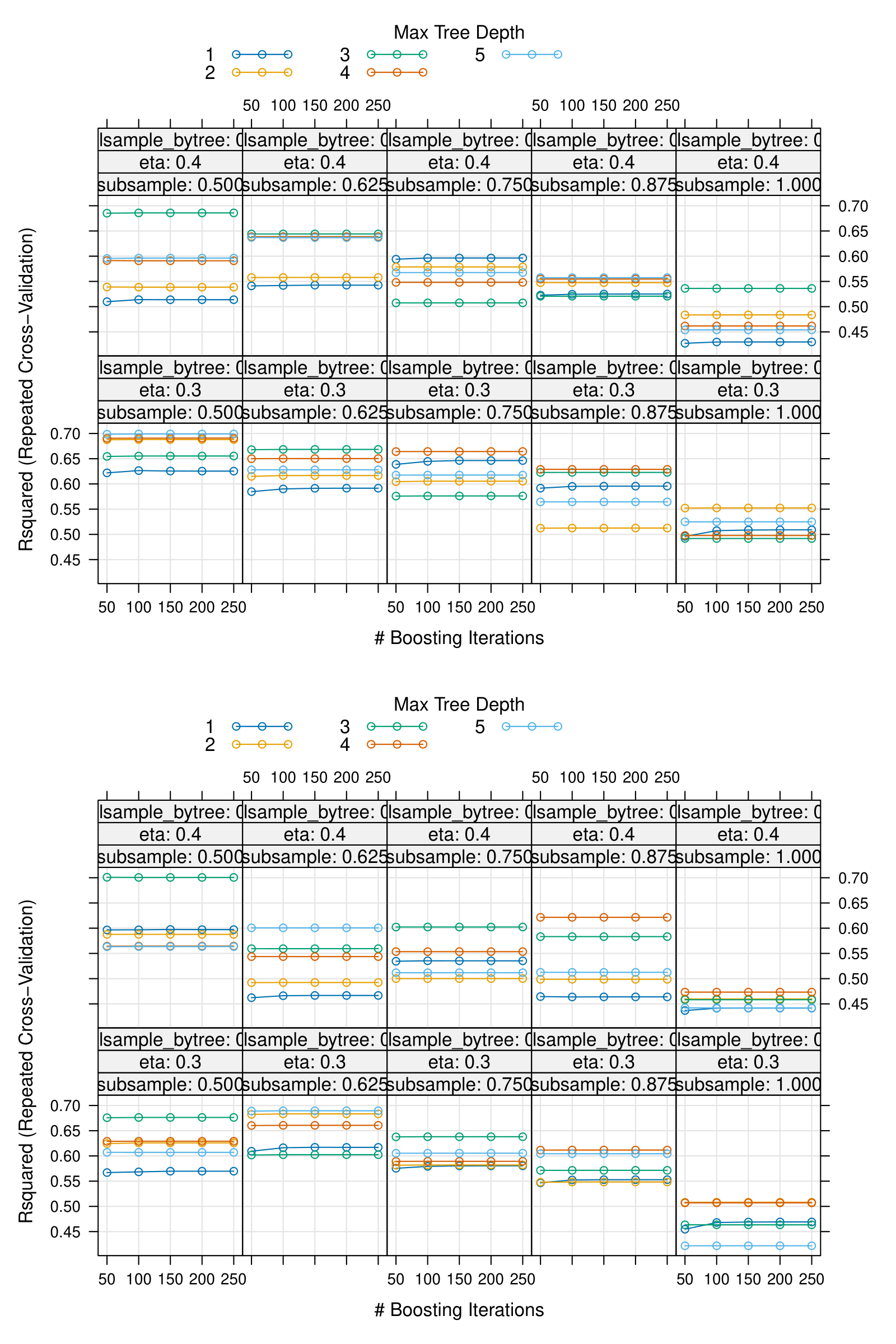
